# Tuberculosis Preventive Therapy for People Living with HIV in Mozambique: Eligibility, Completion, Coverage and TB Disease Rates, April 2021–March 2024

**DOI:** 10.1101/2024.11.25.24317761

**Authors:** Durval Respeito, Yagna Varajidas, José Mizela, Alexandre Nguimfack, Maria Inês Tomo de Deus, Pereira Zindoga, James Cowan, Erica Bila, Benedita Jose, Gisela Sendela, Aleny Couto, Eudoxia Filipe, Lindsay Templin, Sonia Chilundo, Ishani Pathmanathan

## Abstract

**Introduction:** Tuberculosis (TB) is one of the main causes of morbidity and mortality worldwide, especially in people living with HIV (PLHIV). TB preventive treatment (TPT) reduces the incidence and mortality of TB in PLHIV. As of March 2021, in Mozambique, only 40% (182,512/460,080) of those eligible had received a full course of TPT. The aim of this study is to describe the variation in TPT provision and the TB incidence in PLHIV in Mozambique from April 2021 to March 2023.

**Methodology:** We analyzed provincial reports with monthly and semi-annual aggregated data of TPT and TB Monitoring, Evaluation, and Report (MER) indicators of 591 Health Facilities supported by The U.S. Presidential Emergency Plan for Aids Relief (PEPFAR) at the national level. The analyses included the four following indicators: TPT eligibility, TPT completion, TPT coverage, and TB disease incidence rate. TPT data were analyzed using a MS Excel 365 database. Data were collected periodically and analyzed using tables and graphs with variation lines.

**Results:** TPT eligibility decreased by 75%, from 687,711 in April 2021 to 170,011 in March 2024. TPT coverage increased from 42% (489,905/1,177,616) in April 2021 to 89% (1,405,139/1,575,150) in March 2024. TPT completion rate also increased by 8%, from 81% (120,692/148,507) to 89% (104,690/117,764). TPT coverage increased by 47%, from 41% (489.905/1.177.616) in April 2021 to 89% (1.405.139/1.575.150) in March 2024. TB disease incidence rate among PLHIV decreased by 0,15%, from 0.65% (7,801/1,208,559) to 0.5% (7,974/1,592,102).

**Conclusion:** In 3 years, Mozambique drastically increased the number of PLHIV who had received TPT, with almost 90% TPT coverage achieved among PLHIV through a system-wide multidisciplinary approach.

## Introduction

Tuberculosis (TB) is an infectious disease caused by *Mycobacterium tuberculosis (M*.*Tb)*. In 2022, an estimated 10 million people developed active TB globally. TB was the second leading cause of death from a single infectious agent, causing an estimated 1.3 million deaths, leading the list of causes of morbidity and mortality among people living with HIV (PLHIV).^1^

Additionally, a quarter of the global population is estimated to have latent TB Infection (LTBI), defined as a state of persistent immune response to stimulation by *M*.*Tb* antigens with no evidence of clinically active TB.^2^

TB preventive therapy (TPT) can eliminate infection before it progresses to active TB disease. As such, the World Health Organization (WHO) recommends TPT as a service to be integrated in the package of services offered to PLHIV as part of the End TB Strategy until 2030 to reduce TB disease incidence and mortality, especially among those considered at high risk of disease progression, such as PLHIV.^2,3^

Mozambique is heavily affected by both HIV and TB. WHO designates Mozambique as a high-burden country for TB, TB/HIV, and drug-resistant TB.^1^ In 2022, the TB incidence rate was 361 per 100,000 inhabitants, with 110,674 people diagnosed with TB disease and started TB treatment.^4^ In 2023, about 2,166,941 PLHIV in Mozambique were on anti-retroviral therapy (ART), and the TB/HIV co-infection rate was 23.1%.^4,5,6^ Different studies in a similar context to Mozambique have demonstrated three-folder high mortality rate (90 per 100.000 Inh) when compared to TB/HIV co-infected patients.^7,8^

In 2007, the Mozambique Ministry of Health (MISAU) introduced 6 months TPT with Isoniazid (180 doses) to reduce the burden of TB in PLHIV. With the introduction of new HIV instruments in 2019, TPT information among PLHIV was possible to capture.^9^ In 2020, the combined short regimen of Isoniazid and Rifapentine administered once a week for 3 months (12 doses) was introduced.^2,10^ As of March 2021, only 40% (182,512/460,080) of eligible PLHIV had received a full regimen of TPT, either due to missed opportunities at the US or poor adherence and dropout, leaving a gap of 60% (297,568) of those eligible without TPT.^11^

To accelerate reaching PLHIV eligible for TPT, the Mozambique Ministry of Health (MISAU), with PEPFAR’s support, coordinated the implementation of a multidisciplinary strategy at all levels of the health system (national, provincial, district, and Health Facility). This strategy included programmatic prioritization of TPT indicators and inclusion in HIV Quality Improvement cycles (HIV QI), intensive TB screening among PLHIV, identifying PLHIV at-risk for TB, prioritization of shorter and more patient friendly TPT regimens, monitoring adverse events, and using program data to monitor and continuously improve progress toward reaching all eligible PLHIV.^10,12,13^

To understand efforts to scale up TPT, we describe the variation of TPT eligibility, TPT completion, TPT coverage, and reported TB disease incidence among PLHIV in Mozambique from April 2021 to September 2024.

## Methods

This is an observational, descriptive, and retrospective study. Were selected 591 Heath Facilities at the national level. Selection criteria included receiving PEPFAR support during the study period, having an Electronic Patient Tracking System (EPTS), and implementing HIV QI cycles. We used aggregated data of all PLHIV, extracted from EPTS of the HFs. TPT variables analyzed are programmatic indicators: TPT-eligibility, TPT completion (6 months for Isoniazid regimen and 3 months for 3HP short regimen), TPT-coverage, and TB incidence among PLHIV. ^10,12,13^

We analyzed routine programmatic data of PEPFAR indicators (Monitoring, Evaluation, and Reporting)^14^, extracted from EPTS monthly (TPT eligibility and coverage) and semi-annually (TPT completion and TB incidence). Monthly extracted data were from April 2021–March 2024, and semi-annual data from October 2020–March 2021 (FY21Q2) and October 2023–March 2024 (FY24Q2). Data were organized monthly using MS Excel 365 and, descriptive analysis through tables and graphs with variation lines. PLHIV on ART was defined as people with a positive HIV test result enrolled on ART with pharmacy pick-up within 28 days from their last visit.

Systematic screening for TB disease was defined as the identification of people at risk for TB disease in a predetermined target group by assessing symptoms and using diagnostic tests, examinations, or other procedures that can be applied rapidly.^5^ In Mozambique, TB screening is based on the identification of four signs and symptoms of TB (cough, fever, weight loss, and night sweats).^6^ TB screening coverage was defined as the proportion of PLHIV on ART screened for TB among those with a clinical consultation in the semiannual reporting period.

TPT-eligibility was defined as PLHIV on ART who did not have a clear contra-indication such as a positive screening for TB, and who had either not previously received TPT or had been treated for active TB.^10,14^

TPT completion rate was defined as the proportion of ART patients who started a standard course of TPT in the previous reporting period who had documented TPT completion (6 months for INH and, 3 months for 3 HP) based on EPTS recorded TPT medication pickup or clinical review of charts.^10,14^

TPT coverage rate was calculated as the cumulative proportion of PLHIV who completed a course of TPT among TPT-eligible (6 months for INH and, 3 months for 3 HP).^10,14^

The TB disease incidence rate was defined as the number of PLHIV who were diagnosed with active TB and/or started TB treatment among PLHIV on ART.^10,14^

The cascade of reports with aggregated information goes from the US to the national level. These aggregate reports on eligibility, completion, coverage and TB incidence rate are generated in an automated way at the SESP level in the US through specific selection criteria defined in advance.

At the national level, the reports are compiled monthly on a data analysis and visualization platform and are used to measure the performance of HIV programs in preventing progression to active TB disease among people living with HIV (PLHIV).^10,14^ Descriptive statistics (frequencies and percentages) were used to analyze the data. This activity was reviewed by the CDC, deemed not research, and conducted consistent with applicable federal law and CDC policy for non-research ^1^

## Results

In April 2021, the 591 US reported 687,771 PLHIV on ART eligible for TPT, corresponding to 57% (687,771/1,208,559) of the total number of PLHIV on ART in the country (Table 1). By March 2024, this number of those eligible for TPT had fallen by 75% (to 170,011), with a total of 517,700 patients having started TPT.

**Table 1:**
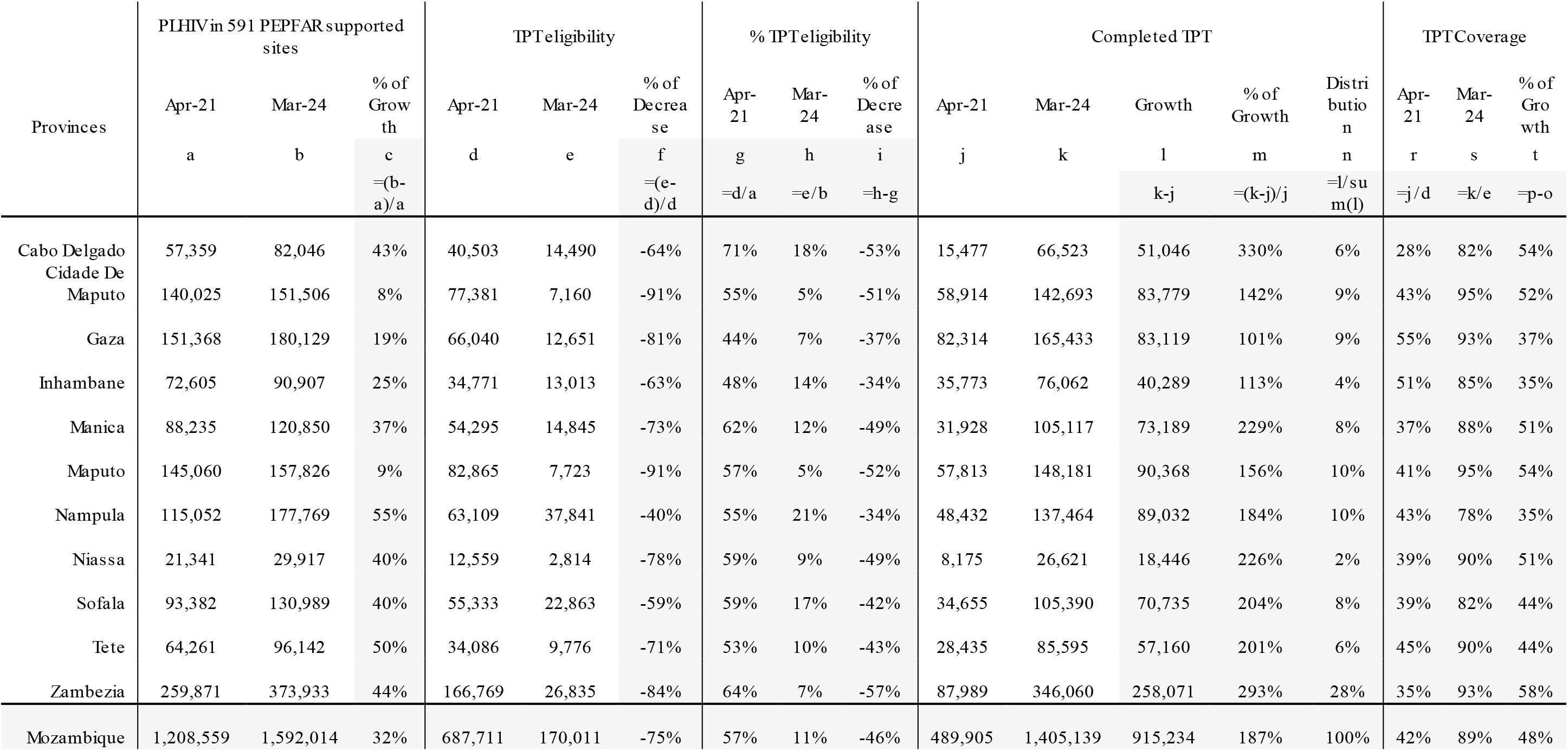
Key Indicators for Tuberculosis Preventive Therapy among People Living with HIV, by province and nationally, April 2021 compared to March 2024.

**Table 2:**
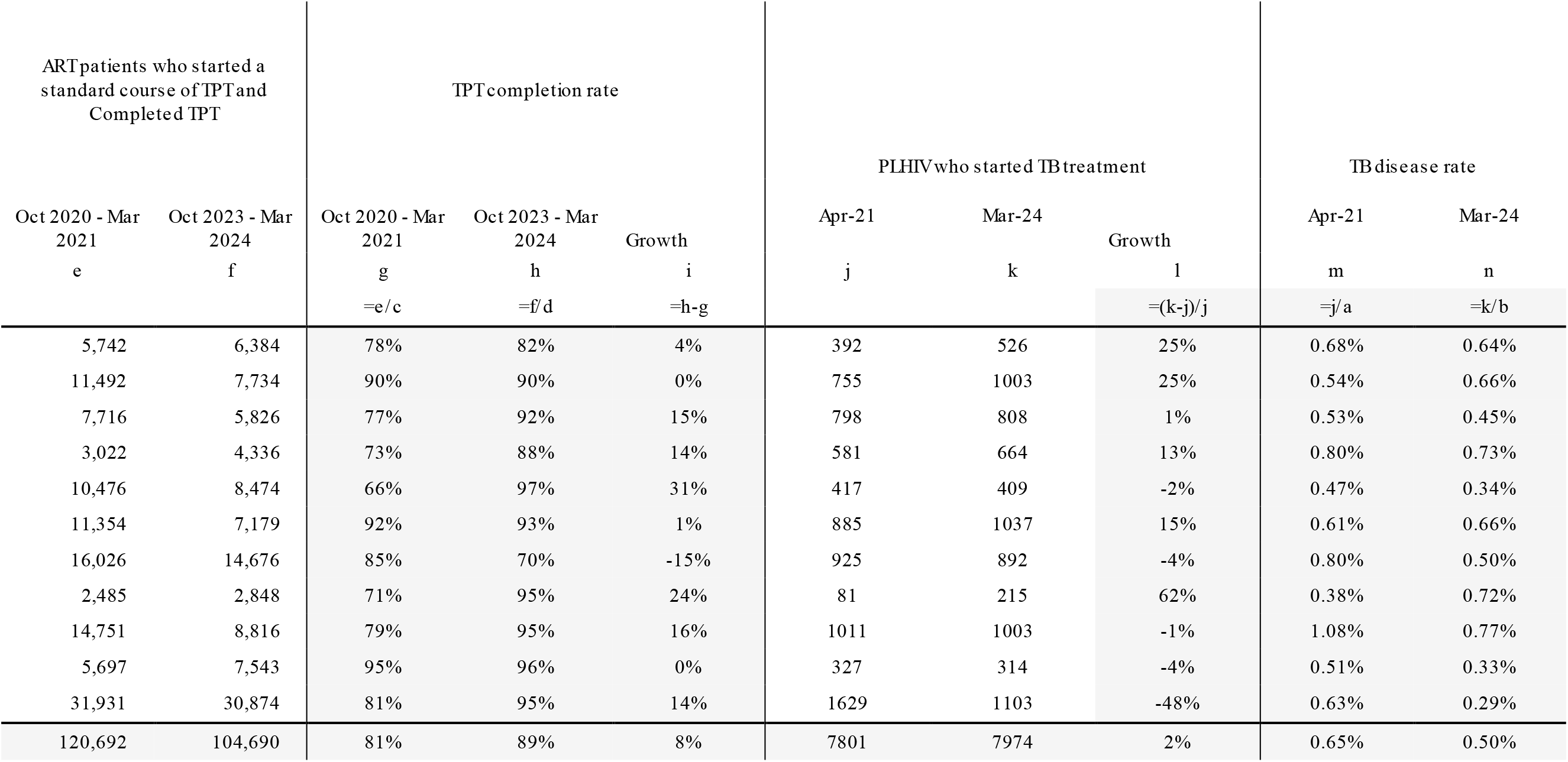
Tuberculosis Preventive Therapy Completion and TB Disease Rates among People Living with HIV, by province and nationally, April 2021 compared to March 2024.

| ART patients who started a standard course of TPT and Completed TPT |  | TPT completion rate |  |  | PLHIV who started TB treatment |  |  | TB disease rate |  |
| --- | --- | --- | --- | --- | --- | --- | --- | --- | --- |
| Oct 2020 - Mar 2021 | Oct 2023 - Mar 2024 | Oct 2020 - Mar 2021 | Oct 2023 - Mar 2024 | Growth | Apr-21 | Mar-24 | Growth | Apr-21 | Mar-24 |
| e | f | g | h | i | j | k | l | m | n |
|  |  | =e/c | =f/d | =h-g |  |  | =(k-j)/j | =j/a | =k/b |
| 5,742 | 6,384 | 78% | 82% | 4% | 392 | 526 | 25% | 0.68% | 0.64% |
| 11,492 | 7,734 | 90% | 90% | 0% | 755 | 1003 | 25% | 0.54% | 0.66% |
| 7,716 | 5,826 | 77% | 92% | 15% | 798 | 808 | 1% | 0.53% | 0.45% |
| 3,022 | 4,336 | 73% | 88% | 14% | 581 | 664 | 13% | 0.80% | 0.73% |
| 10,476 | 8,474 | 66% | 97% | 31% | 417 | 409 | -2% | 0.47% | 0.34% |
| 11,354 | 7,179 | 92% | 93% | 1% | 885 | 1037 | 15% | 0.61% | 0.66% |
| 16,026 | 14,676 | 85% | 70% | -15% | 925 | 892 | -4% | 0.80% | 0.50% |
| 2,485 | 2,848 | 71% | 95% | 24% | 81 | 215 | 62% | 0.38% | 0.72% |
| 14,751 | 8,816 | 79% | 95% | 16% | 1011 | 1003 | -1% | 1.08% | 0.77% |
| 5,697 | 7,543 | 95% | 96% | 0% | 327 | 314 | -4% | 0.51% | 0.33% |
| 31,931 | 30,874 | 81% | 95% | 14% | 1629 | 1103 | -48% | 0.63% | 0.29% |
| 120,692 | 104,690 | 81% | 89% | 8% | 7801 | 7974 | 2% | 0.65% | 0.50% |

The downward trend was rapid until January 2022, with a slight increase in those eligible until May 2022 and then a decrease again until January 2023, and then stabilization in a straight line and minimal variations until March 2024. In proportional terms, all provinces saw a reduction in those eligible for the TPT, with Maputo and Maputo City provinces standing out with the biggest reduction, both at 91%, followed by Zambézia province, which saw a reduction of 84%. On the other hand, Nampula province saw a smaller reduction of 40%.

TPT completion increased by 915,234, from 489,905 (41%) in April 2021 to 1,405,139 (88%) in March 2024, representing an increase of 187%. The provinces of Cabo Delgado and Zambézia (the province with the highest number of PLHIV on ART) saw the highest growth in the cumulative number of PLHIV completing TPT, with 330% and 293%, respectively. The lowest growth was recorded in Gaza province, with 101%.

TPT coverage increased from 42% (489,905/1,177,616) in April 2021 to 89% (1,405,139/1,575,150) in March 2024, with almost all provinces above 80%, except Nampula province (Table 1; Graph).

TPT coverage also increased by 48%, from 42% to 89%. The trend in all the provinces was positive, and in September 2024, coverage was above 80%, except for Nampula province, which had 78% coverage. The provinces of Maputo City (95%), Maputo Province (95%), Gaza (93%), Zambézia (93%), Tete (90%) and Niassa (90%) had the best performance.

From March 2021 to March 2024, although the number of PLHIV starting TB treatment increased by 2%, from 7,801 to 7,974, the reported TB incidence rate among PLHIV decreased from 0.65% (7,801/1,208,559) in 2021 to 0.5% (7,974/1,592,102) in 2024. At the national level, the provinces of Zambézia, Sofala, and Nampula stand out with a reduction in the TB incidence rate of 0.33%, 0.32%, and 0.3%, respectively. On the other hand, Maputo City and Niassa saw an increase in the incidence rate, with 0.12% and 0.34%.

## Discussion

The multidisciplinary strategy coordinated by the MoH since April 2021 has been crucial to achieving the current gains in PLHIV TPT in the country. Despite the gradual reduction in eligibility, there has been a proportional increase in initiation, completion and coverage of TPT in PLHIV. These gains have been achieved by making integrated use of the available resources, which include not only human resources, but also strategic information systems that guide interventions based on data.

In April 2021, there was a 60% gap between those eligible for TPT and those who had received a full TPT course, possibly related to the loss of opportunities to start TPT. There was an increase in those eligible between January and May 2022. This increase could be associated with weaknesses inherent in the Health System (stock-outs of TPT regimens or lack of updating of the SESP or increase in eligibility in Nampula), the care provider (greater identification of eligibles with missed opportunities) and the patient (insufficient literacy about TPT, missed follow-ups).^15^

TPT completion and coverage are two factors that depend mainly on the adherence of PLHIV to TPT. These indicators remained below 85%, as in the provinces of Nampula (72%), Cabo Delgado (82%) and Sofala (82%). The gap between those eligible and those who have started TPT has narrowed by 48%, from 60% in April 2021 to 12% in September 2024. In 16 African countries supported by PEPFAR (South Africa, Tanzania, Kenya, Lesotho, Tanzania, among others) report a similar experience, with low performance of TPT indicators, followed by coverage above 80%.^16,17,21^

Targeted interventions to increase TPT completion and coverage in low-performing provinces include identifying sites and geographic areas with low TPT performance for intensive follow-up and mentoring; sensitizing all patients on ART and consistently offering TPT among newly enrolled users; differentiated TPT dispensing options (e.g. multi-monthly TPT dispensing); and adherence counseling, psychosocial support and monthly monitoring for users on TPT.

The recent introduction of short-term TPT in the 4 provinces of the southern region of the country and its expansion throughout the country is an opportunity to improve TPT adherence and completion. Additionally, there are provincial variations that may require adapted interventions. For example, in Cabo Delgado province, the ongoing military conflict since 2018 has resulted in US closures and population displacement (including to Nampula province), resulting in difficult access for TPT In this province, TPT is offered through mobile clinics and brigades in resettlement and accommodation centers and through military providers.^18^

Finally, the positive impact of this huge expansion of TPT is reflected in the reduction of the reported TB incidence rate by 0.15% (from 0.65% to 0.5%), especially in the context of a higher estimated number of PLHIV and a better scale and quality of TB screening among them However, there are still challenges in the quality of TB screening due to the subjectivity of the four signs and symptoms in PLHIV, and the number of PLHIV diagnosed with TB among PLHIV in general increased by 2% (from 7,801 to 7,974) during the reference periods.^19^

Mozambique’s HIV and TB programs are working to respond to the reduction in the incidence rate through periodic training of professionals with clinic sessions to improve the quality of TB symptom screening and referral by providers; updating TB screening guidelines with the inclusion of chest X-ray and computer-assisted diagnosis in US where it is available and the expansion of the short TPT regimen to the whole country.^20^ The options available for rapid TB diagnosis are also increasing, including TB LAM assays, TrueNat and GeneXpert. On the other hand, the implementation of infection control measures in the US and in communities is fundamental for TB control.

This study has several limitations. Although the PEPFAR-supported sites included in this study represent 80% of PLHIV on ART in Mozambique, the performance of the TPT program may be different in sites that do not receive direct PEPFAR support. As secondary program data was used, it was not possible to analyze or evaluate individual patient records. In addition, PLHIV can access health services, including TPT, in any location, regardless of where they live, so some people may have been counted more than once.

TPT coverage among PLHIV in Mozambique increased dramatically between 2021 and 2024 to reach 89% of estimated eligible PLHIV. To sustain and accelerate the TPT response, the MoH and partners continue to work to improve access to quality TPT services for PLHIV still eligible for TPT and/or newly enrolled on ART, to ultimately reduce TB-related morbidity and mortality among PLHIV in the country.

## Conclusion

In 3 years, Mozambique has dramatically increased the number of PLHIV who have received TPT and completed it, as well as their coverage. As a result, around 90% TPT coverage was achieved among this large cohort of PLHIV. We hope that **TB incidence rates** among PLHIV will continue to reduce as observed in this study, as ART and TPT coverage continues to increase, and Mozambique moves closer to controlling the HIV epidemic.

## Data Availability

All data produced are available from PEPFAR Monitoring, Evaluation, and Reporting program data.

https://data.pepfar.gov/datasets#PDD

## Acknowledgements

We would like to acknowledge the Mozambique Ministry of Health, PEPFAR Implementing Partners and all those contributed to this work.

## Funding Acknowledgement

This manuscript has been supported by the President’s Emergency Plan for AIDS Relief (PEPFAR), through the Centers for Disease Control and Prevention (CDC) under the terms of *CGH* HRS2017-519, CGH HSR2441, P-2017-008 and P-2019-008. The findings and conclusions in this manuscript are those of the author(s) and do not necessarily represent the official position of the funding agencies (CDC, OGAC/PEPFAR).

---

**Figure 1:**
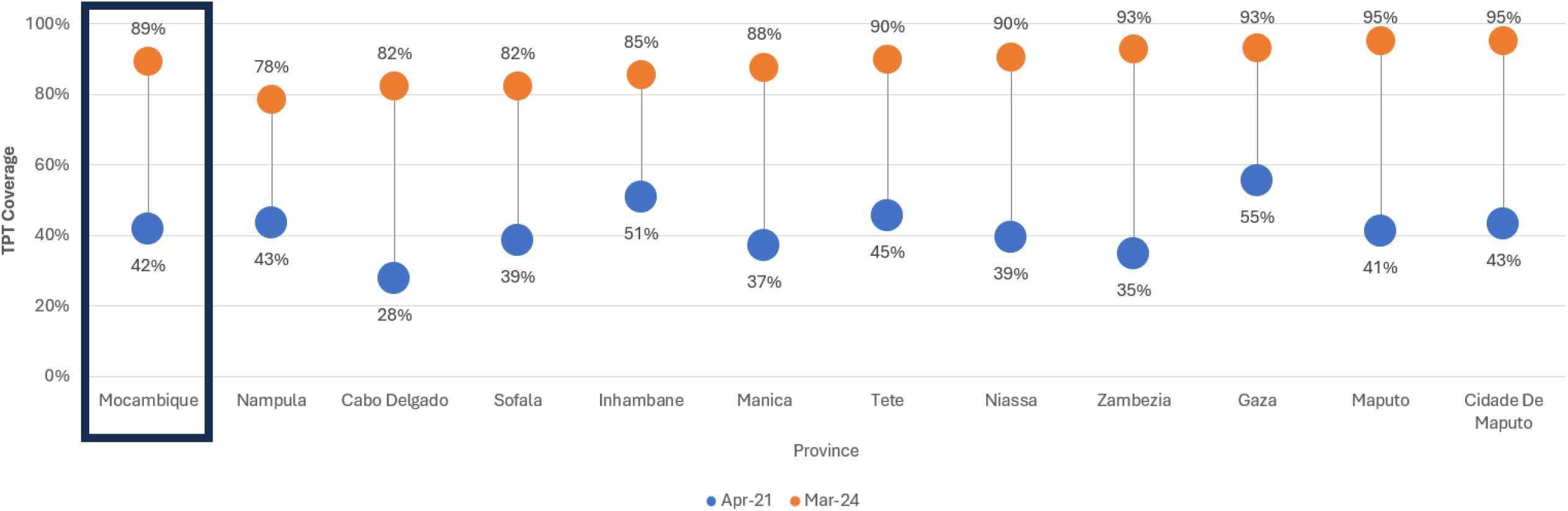
Change in TB Preventive Therapy Coverage Rates among People Living with HIV, by Provinces, April 2021 to March 2024.

## Footnotes

1 **\***45 C.F.R. part 46.102(l)(2), 21 C.F.R. part 56; 42 U.S.C. Sect. 241(d); 5 U.S.C. Sect. 552a; 44 U.S.C. Sect. 3501 et seq

